# QRS 3D Voltage-Time Integral in Narrow QRS Complex - Establishing the Normal Reference Range

**DOI:** 10.1101/2025.02.12.25322179

**Authors:** Amulya Gupta, Christopher J. Harvey, Uzair Mahmood, Jacob D. Baer, Nikhil Parimi, Ashutosh Bapat, Seth H. Sheldon, Madhu Reddy, Zijun Yao, Yongkuk Lee, Amit Noheria

## Abstract

**Background:** Vectorcardiographic 3D QRS voltage-time integral (VTI_QRS-3D_) is a novel marker of ventricular dyssynchrony pertinent for cardiac resynchronization therapy. It may have additional clinical utility but its normal reference ranges have not been established. We sought to define reference ranges for VTI_QRS-3D_ in healthy individuals.

**Methods:** We retrospectively analyzed 12-lead ECGs of healthy adults (2010-2014) and compared them to patients with cardiomyopathy with reduced ejection fraction (EF) <50%. Using the Kors matrix, 12-lead ECGs with QRS duration ≤120 ms were converted to vectorcardiographic X, Y, and Z leads. VTI_QRS-3D_ was calculated as the instantaneous root-mean-square (3D) voltage integrated over the QRS duration. Reference range limits were defined as the 2.5th to 97.5th percentiles respectively for healthy females and males in age groups 18-34, 35-54 and ≥55 years.

**Results:** The study included 468 healthy adults (age 44.6 ± 17.0 years; 63.9% female) and 314 patients with cardiomyopathy (age 62.1 ± 14.0 years; 34.4% female). VTI_QRS-3D_ was significantly larger in the cardiomyopathy patients compared to the healthy population (48.2±21.4 vs. 38.1±9.3 µVs, p<0.0001). Increased age and female sex were significant predictors of lower VTI_QRS-3D_ in the healthy population (both p<0.0001). VTI_QRS-3D_ reference ranges for respective age groups for healthy females were 23.2-55.0, 23.9-56.4 and 19.6-50.9 µVs, and for healthy males were 29.9-57.2, 28.2-56.7 and 21.4-55.9 µVs.

**Conclusion:** VTI_QRS-3D_ is higher at younger age in healthy population, male sex and in patients having cardiomyopathy with reduced EF. Age and sex need to be accounted for using VTI_QRS-3D_ as a marker for structural heart disease.

## INTRODUCTION

Electrocardiograms (ECGs) are recorded using a standard 12-lead configuration, which includes 3 limb and 6 precordial electrodes. While this 12-lead ECG format is widely used and supported by over a century of research, it represents cardiac electrical activity along anatomically arbitrary axes rather than providing a true three-dimensional (3D) depiction of cardiac electrical activity. Vectorcardiography (VCG) addresses this limitation by representing cardiac electrical activity along the orthogonal cartesian axes X (right-to-left), Y (cranial-to-caudal), and Z (anterior-to-posterior).^1^ Although VCG is seldom performed in contemporary clinical practice, it can be derived from a 12-lead ECG using various transformation matrices such as Kors’s or Dower’s regression matrices.^2^ Plotting the root-mean-square (RMS) of the instantaneous voltages of X, Y, and Z leads yields the 3D ECG, which is a scalar representation of the net surface cardiac voltages (**Figure 1**).

**Figure 1.**
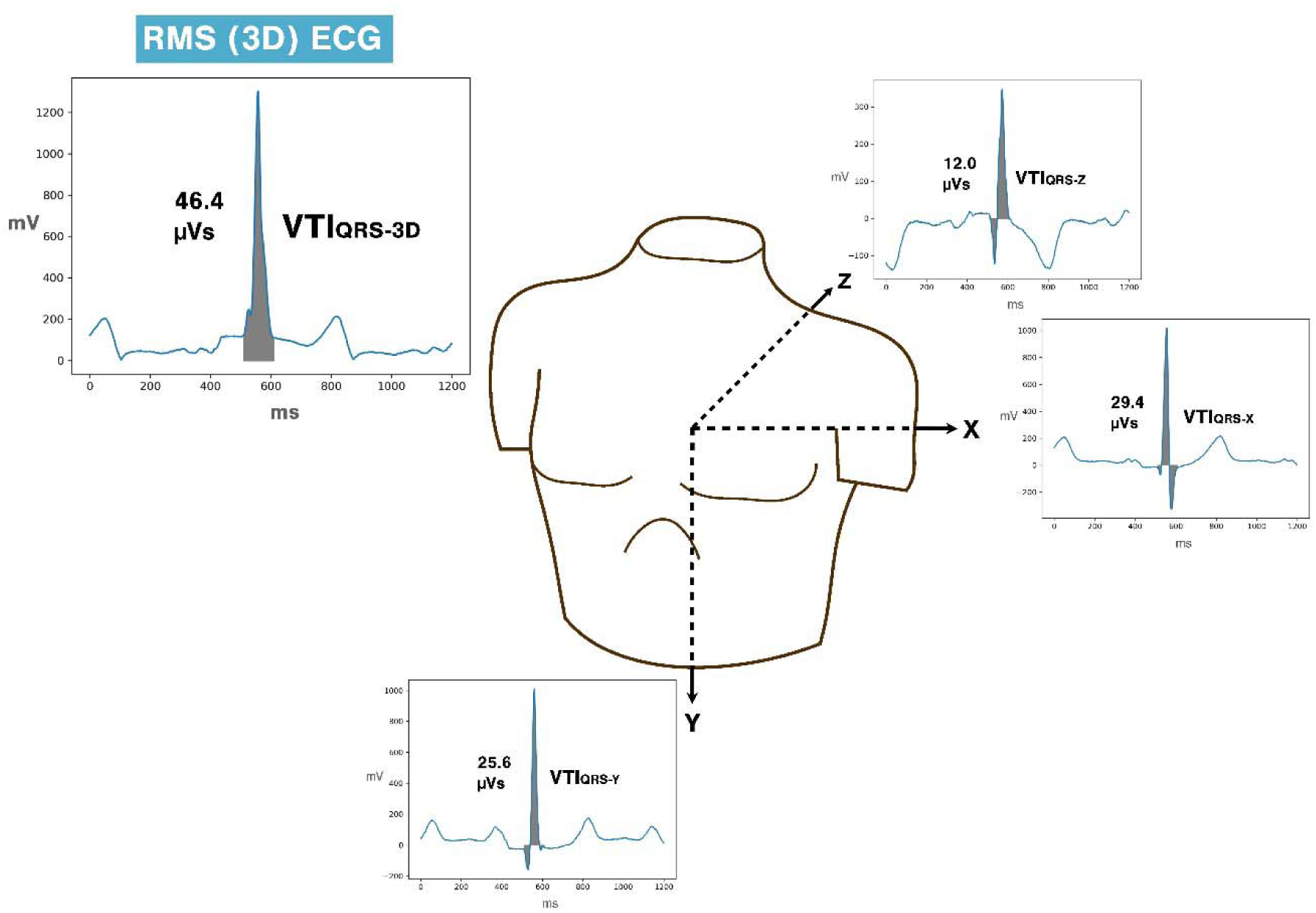
Schematic representation of VTI_QRS-3D_ calculation.

Recent work has established voltage-time integral of the 3D ECG QRS complex (VTI_QRS-3D_) and a related metric, 3D-QRS area, as novel markers of left ventricular electrical dyssynchrony.^3,4^ By integrating the instantaneous 3D voltage over the duration of QRS complex, VTI_QRS-3D_ quantifies the total ECG potentials recorded during ventricular depolarization. Mechanistically, electrical dyssynchrony fragments the depolarization wavefront, disrupting the cancellation of opposing synchronized wavelets and leads to increased electrical potentials which manifests as higher VTI_QRS-3D_.^5,6^ VTI_QRS-3D_ has proven to be a stronger predictor of response to cardiac resynchronization therapy (CRT) compared to QRS duration.^4^ Its utility extends beyond CRT response, as previous studies have also demonstrated the application of VTI_QRS-3D_ in identifying left ventricular hypertrophy.^7,8^

Despite its advantages, VTI_QRS-3D_ is not routinely reported in clinical ECGs, and limited literature exists regarding its normal range beyond the contexts of CRT and left ventricular hypertrophy. This study aims to establish a reference range for VTI_QRS-3D_ in healthy patients without cardiac electrical or structural disease. Specifically, we evaluated VTI_QRS-3D_ in healthy individuals with normal ECGs and narrow QRS complexes, and compared to patients having cardiomyopathy with reduced EF and normal QRS duration.

## METHODS

This study was conducted under approval by the Institutional Review Board at The University of Kansas. We conducted a retrospective analysis on ECGs from 2010-2014 at The University of Kansas Medical Center (KUMC). This retrospective cohort consisted of two populations with narrow QRS complexes (≤120 ms). The first population, referred to as ‘healthy group’, consisted of individuals lacking a history of cardiomyopathy and ECG conduction abnormalities while the second population, referred to as ‘cardiomyopathy group’, consisted of patients with a known diagnosis of cardiomyopathy with reduced EF and a narrow QRS complex.

The healthy population was queried using Healthcare Enterprise Repository for Ontological Narration (HERON), which is a repository of all health visit International Classification of Diseases (ICD) codes combined with a variety of hospital and medical center electronic records.^9,10^ We identified patients with an outpatient routine preventative health visit code and a procedure code for ECG between 2010 and 2014. We excluded patients with diagnostic codes for any cardiovascular disease or chronic non-communicable disease diagnosis. We then manually downloaded their digital ECG files in .xml and .pdf formats from the Philips® IntelliSpace^TM^ ECG management system. All ECGs with QRS duration >120 ms were excluded. The ECGs were then reviewed by an experienced electrophysiologist (AN) for any abnormal findings. Patient charts were then manually reviewed to identify any cardiovascular or physical disease condition, upon identification of which these ECGs were also excluded.

The cardiomyopathy with reduced EF patients were also queried using HERON for echocardiographic left ventricular ejection fraction below 50%. In this group, patients with a history of cardiac arrhythmias or conduction abnormalities were excluded. Clinical echocardiographic reports were used to extract left ventricular dimensions and ejection fraction which were measured according to the American Society of Echocardiography guidelines.^11^

### ECG processing

Clinical 12-lead ECG .xml files were retrieved from the Philips® IntelliSpace^TM^ ECG management system and processed using Python. The 12-lead 1200 ms representative beat ECG signals were converted to orthogonal X, Y, Z leads using the Kors conversion matrix.^12^ RMS of the orthogonal leads was computed to generate a 3D ECG signal. The location of QRS onset and QRS duration were obtained from the proprietary Philips DXL algorithm. VTI_QRS-3D_ was obtained by integrating the voltage across the QRS complex. Similarly, individual VTI_QRS_ for X, Y, Z leads were also calculated (**Figure 1**).

### Statistical analysis

Continuous variables were expressed as mean ± standard deviation (SD) and categorical variables as n (%). VTI_QRS-3D_ values were reported using mean ± SD, median and percentiles (2.5^th^, 25^th^, 75^th^ and 97.5^th^). Reference ranges were defined as values between 2.5^th^ and 97.5^th^ percentiles. Continuous variables were compared using independent sample t-test and one way ANOVA, and categorical variables were compared using the χ2-squared test. Univariate and multivariate linear regression was used to assess association between predictor variables and VTI_QRS-3D_, with results expressed as a β-coefficient ± standard error (SE). All statistical analyses were done in JMP Pro 17 (SAS Inst. Cary, NC, USA) and R (R version 4.4.1).

## RESULTS

The healthy group included 468 adults. The QRS duration in this healthy population was 86.9 ± 9.7 ms. The VTI_QRS_ in the vectorcardiographic X, Y, Z leads was 23.8 ± 7.1 µVs, 19.1 ± 7.7 µVs and 15.4 ± 7.1 µVs respectively. The VTI_QRS-3D_ among this healthy group was 38.1 ± 9.3 µVs. The cardiomyopathy with reduced EF group included 314 patients. The baseline demographic, echocardiographic and ECG variables for both groups are summarized in **Table 1**.

**Table 1.** Baseline characteristics in the normal and cardiomyopathy with reduced EF populations.

| **Variables** | **Healthy group**<br>**(n=468)** | **Cardiomyopathy group**<br>**(n=314)** | **p-value** |
| --- | --- | --- | --- |
| **Demographics** |  |  |  |
| Age, years | 44.6 ± 17.0 | 62.1 ± 14.0 | **<0.0001** |
| Women | 299 (63.9%) | 108 (34.4%) | **<0.0001** |
| Race |  |  | **0.001** |
| White | 312 (66.7%) | 197 (62.7%) |  |
| Black | 73 (15.6%) | 79 (25.2%) |  |
| Other | 83 (17.7%) | 38 (12.1%) |  |
| Body surface area, m<sup>2</sup> | 1.85 ± 0.27 | 1.98 ± 0.31 | **<0.0001** |
| **Echocardiogram** | *n=132* |  |  |
| LV internal dimension in diastole, cm | 4.4 ± 0.6 | 5.3 ± 0.8 | **<0.0001** |
| LV internal dimension in systole, cm | 2.9 ± 0.5 | 4.2 ± 0.9 | **<0.0001** |
| LV ejection fraction, % | 59.5 ± 3.5 | 35.9 ± 9.4 | **<0.0001** |
| LV mass, g | 127.1 ± 38.6 | 219.6 ± 79.9 | **<0.0001** |
| LV mass indexed, g/m<sup>2</sup> | 68.8 ± 16.6 | 110.6 ± 36.1 | **<0.0001** |
| Interventricular septum, cm | 0.9 ± 0.2 | 1.1 ± 0.2 | **<0.0001** |
| LV posterior wall, cm | 0.9 ± 0.2 | 1.1 ± 0.2 | **<0.0001** |
| **ECG** |  |  |  |
| QRS duration, ms | 86.9 ± 9.7 | 94.1 ± 11.4 | **<0.0001** |
| Amplitude<sub>QRS-3D</sub>, mV | 1.27 ± 0.37 | 1.34 ± 0.62 | **0.03** |
| VTI<sub>QRS-3D</sub>, μVs | 38.1 ± 9.3 | 48.2 ± 21.4 | **<0.0001** |
| VTI<sub>QRS-X</sub>, μVs | 23.8 ± 7.1 | 28.0 ± 16.0 | **<0.0001** |
| VTI<sub>QRS-Y</sub>, μVs | 19.1 ± 7.7 | 19.5 ± 12.2 | **0.6** |
| VTI<sub>QRS-Z</sub>, μVs | 15.4 ± 7.1 | 25.7 ± 14.6 | **<0.0001** |

### Baseline demographics

In the healthy group, 299 (63.9%) were female, with a mean age of 44.6 ± 17.0 years, and 312 (66.7%) identified as white. In the cardiomyopathy group, 108 (34.4%) were female, the mean age was 62.1 ± 14.0 years, and 197 (62.7%) identified as white. All the baseline demographic variables differed significantly between the groups (all p <0.05).

### Echocardiogram variables

Echocardiographic variables were available for 132 (27.8%) patients in the healthy group, and all patients in the cardiomyopathy group. The healthy group demonstrated smaller left ventricular internal dimensions in diastole (LVIDd: 4.4 ± 0.6 cm vs. 5.3 ± 0.8 cm) and systole (LVIDs: 2.9 ± 0.5 cm vs. 4.2 ± 0.9 cm) compared to the cardiomyopathy group (both p <0.0001). Left ventricular ejection fraction (LVEF) was higher in the healthy group (59.5 ± 3.5 vs. 35.9 ± 9.4, p <0.0001). Additionally, the healthy group had lower left ventricular mass indexed (LVMi: 68.8 ± 16.6 g/m^2^ vs. 110.6 ± 36.1 g/m^2^, p <0.0001), interventricular septum thickness (0.9 ± 0.2 cm vs. 1.1 ± 0.2 cm, p <0.0001), and left ventricular posterior wall thickness (0.9 ± 0.2 cm vs. 1.1 ± 0.2 cm, p <0.0001).

### ECG variables

The healthy group had shorter QRS duration compared to the cardiomyopathy group (86.9 ± 9.7 ms vs. 94.1 ± 11.4 ms, p <0.0001). Among the VCG variables, the healthy group had smaller Amplitude_QRS-3D_ (1.27 ± 0.37 mV vs. 1.34 ± 0.62 mV, p =0.03), VTI_QRS-X_ (23.8 ± 7.1 µVs vs. 28.0 ± 16.0 µVs, p <0.0001), VTI_QRS-Z_ (15.4 ± 7.1 µVs vs. 25.7 ± 14.6 µVs, p <0.0001), and VTI_QRS-3D_ (38.1 ± 9.3 µVs vs. 48.2 ± 21.4 µVs, p <0.0001). VTI_QRS-Y_ was similar between both groups (19.1 ± 7.7 µVs vs. 19.5 ± 12.2 µVs, p =0.6).

### Vectorcardiographic lead VTI _QRS_ in healthy group

The distribution of VTI_QRS_ for vectorcardiographic X, Y and Z leads for the healthy group are shown in **Table 2A-B.** The VTI_QRS_ values in X and Z leads are smaller for females compared to males (both p <0.0001), while VTI_QRS-Y_ shows no sex-based difference (p =0.8). In general, the VTI_QRS_ for Y (p <0.0001) and X (p =0.02) leads decreases with increasing age, while VTI_QRS-Z_ does not show any age-related trend.

**Table 2A.** Distributions of VTI_QRS_ in vectorcardiographic X, Y, Z leads in healthy females (n=299)

| Variable | Mean ± S.D. (Healthy females) |  |  |
| --- | --- | --- | --- |
| VTI<sub>QRS</sub>, μVs | X | Y | Z |
| **Overall** | 22.0 ± 6.1 | 19.0 ± 7.6 | 14.0 ± 6.4 |
| **Age, years** | **p = 0.02** | **p < 0.0001** | p = 0.9 |
| 18-34 | 22.8 ± 5.7 | 21.7 ± 7.9 | 14.0 ± 6.6 |
| 35-54 | 22.7 ± 6.8 | 18.8 ± 7.2 | 13.9 ± 6.9 |
| 55-64 | 20.6 ± 4.9 | 16.9 ± 6.8 | 13.6 ± 5.6 |
| ≥65 | 20.2 ± 6.2 | 16.4 ± 7.4 | 14.6 ± 6.1 |
| **Race** | p = 0.3 | **p = 0.04** | p = 0.3 |
| White | 21.9 ± 6.3 | 18.6 ± 7.4 | 13.9 ± 6.3 |
| Black | 23.1 ± 5.9 | 21.3 ± 8.6 | 13.3 ± 6.0 |
| Other/unknown | 21.2 ± 5.3 | 17.9 ± 6.7 | 15.2 ± 7.3 |
| **Body Composition** | **β ± SE (p-value)** |  |  |
| Body surface area, m<sup>2</sup> | 2.0 ± 1.6<br>(p = 0.2) | -1.4 ± 2.0<br>(p = 0.5) | 2.4 ± 1.6<br>(p = 0.1) |
| Body mass index, kg/m<sup>2</sup> | -0.05 ± 0.06<br>(p = 0.4) | -0.18 ± 0.08<br>**(p = 0.02)** | 0.06 ± 0.06<br>(p = 0.3) |

**Table 2B.** Distributions of VTI_QRS_ in vectorcardiographic X, Y, Z leads in healthy males (n=169)

| Variable | Mean ± S.D. (Healthy males) |  |  |
| --- | --- | --- | --- |
| VTI<sub>QRS</sub>, μVs | X | Y | Z |
| **Overall** | 26.9 ± 7.5 | 19.2 ± 8.0 | 18.0 ± 7.5 |
| **Age, years** | **p = 0.02** | **p < 0.0001** | p = 0.09 |
| 18-34 | 29.0 ± 6.9 | 23.0 ± 7.3 | 18.6 ± 7.9 |
| 35-54 | 26.7 ± 7.1 | 18.2 ± 7.6 | 19.2 ± 7.8 |
| 55-64 | 23.8 ± 8.3 | 16.4 ± 7.7 | 15.3 ± 5.7 |
| ≥65 | 26.2 ± 7.6 | 15.5 ± 7.4 | 16.6 ± 7.1 |
| **Race** | p = 0.5 | p = 0.8 | p = 0.3 |
| White | 26.8 ± 7.2 | 19.0 ± 8.2 | 17.5 ± 7.7 |
| Black | 28.8 ± 7.5 | 20.4 ± 5.3 | 17.7 ± 7.1 |
| Other/unknown | 26.3 ± 8.3 | 19.2 ± 8.6 | 19.6 ± 7.3 |
| **Body Composition** | **β ± SE (p-value)** |  |  |
| Body surface area, m<sup>2</sup> | 1.0 ± 2.0<br>(p = 0.6) | -4.9 ± 2.1<br>**(p = 0.02)** | 3.5 ± 2.0<br>(p = 0.09) |
| Body mass index, kg/m<sup>2</sup> | -0.05 ± 0.11<br>(p = 0.6) | -0.32 ± 0.12<br>**(p = 0.007)** | -0.16 ± 0.11<br>(p = 0.2) |

### 3D RMS ECG VTI _QRS_ in healthy group

The distribution of VTI_QRS-3D_ among different demographic categories are summarized in **Table 3A** for females and **Table 3B** for females. Briefly, VTI_QRS-3D_ showed a decreasing trend across age groups from 18 to 65 years (p =0.003 for females, p <0.0001 for males). The values of VTI_QRS-3D_ were similar across racial groups (p =0.2 females, p =0.5 males), and did not exhibit statistically significant association with body surface area (BSA, p=0.5 females, p=0.08 males) or body mass index (BMI, p=0.2 females, p=0.08 males). The distribution of VTI_QRS-3D_ across age groups for healthy population is summarized in **Figure 2**.

**Figure 2.**
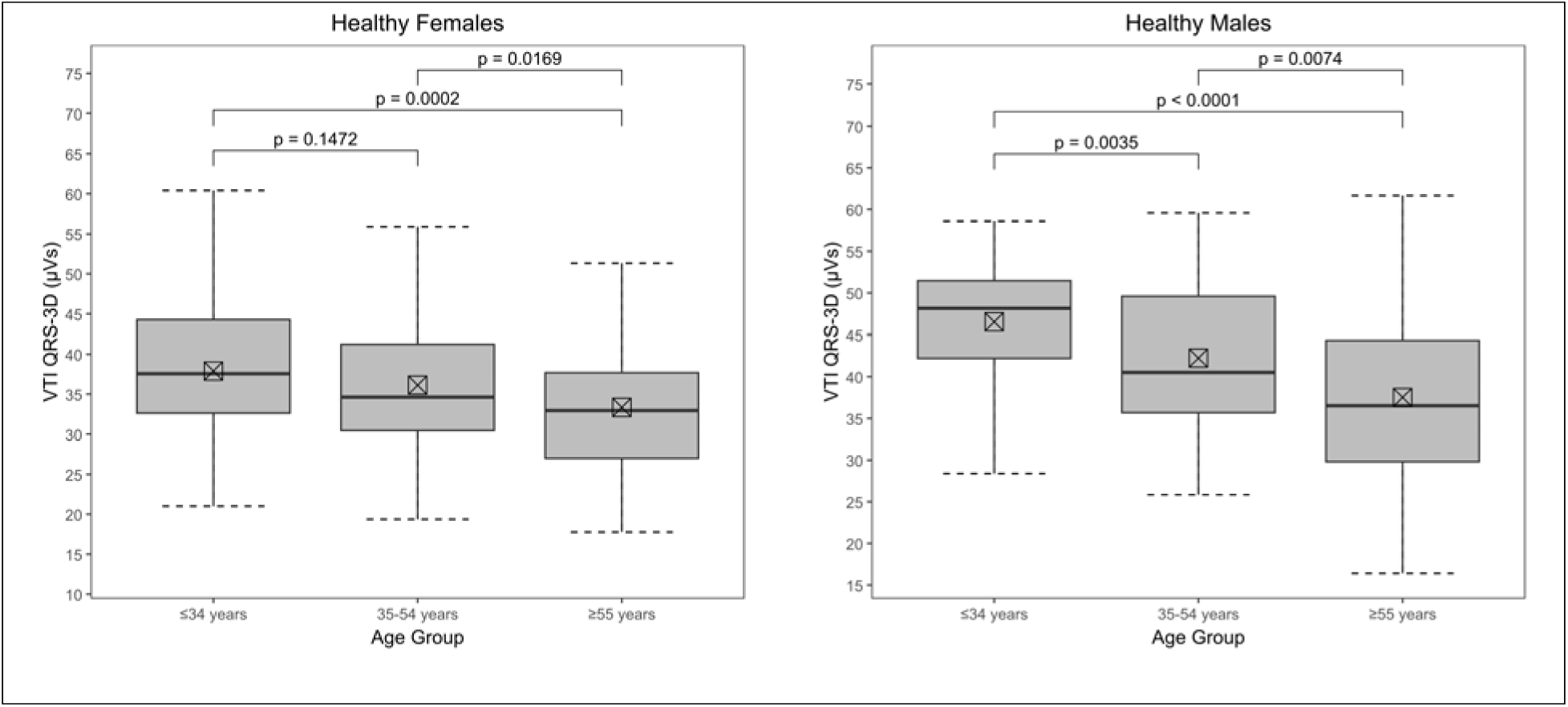
Boxplots for VTI_QRS-3D_ by age groups for healthy females (n=299) and healthy males (n=169) ⍰ indicates mean; horizontal line indicates median; p-values shown for unpaired t-test

**Table 3A.** Distribution of VTI_QRS-3D_ among healthy females.

| **Variables**<br>**(Healthy females)** | **n** | **VTI<sub>QRS-3D</sub>, $\mu\text{Vs}$**<br>**(Mean ± SD)** | **IQR** | **Reference ranges**<br>**2.5<sup>th</sup>-97.5<sup>th</sup>**<br>**percentile** | **p-value** |
| --- | --- | --- | --- | --- | --- |
| **Overall** | 299 | 35.7 ± 8.6 | 29.6-41.4 | 20.2-55.7 | - |
| **Age, years** |  |  |  |  | **0.003** |
| 18-34 | 92 | 37.9 ± 8.5 | 32.6-44.3 | 23.2-55.0 | |
| 35-54 | 107 | 36.1 ± 8.6 | 30.5-41.2 | 23.9-56.4 | |
| 55-64 | 58 | 33.3 ± 7.5 | 28.2-36.9 | 22.3-51.0 | |
| $\geq 65$ | 42 | 33.4 ± 8.9 | 25.5-39.3 | 19.1-46.8 | |
| **Race** |  |  |  |  | 0.2 |
| White | 201 | 35.3 ± 8.6 | 29.6-40.7 | 20.1-54.6 | |
| Black | 53 | 37.6 ± 8.9 | 30.8-43.1 | 24.5-55.7 | |
| Other/unknown | 45 | 35.4 ± 8.2 | 30.1-41.6 | 21.2-49.6 | |
| **Body Composition** |  | **Mean ± SD** | **β-**<br>**coefficient** |  |  |
| Body surface area, m<sup>2</sup> | 298 | 1.8 ± 0.2 | -1.5 ± 2.2 | | 0.5 |
| Body mass index, kg/m<sup>2</sup> | 298 | 26.1 ± 5.8 | -0.1 ± 0.1 | | 0.2 |
| **Echocardiography** |  | **Mean ± SD** | **β-**<br>**coefficient** |  |  |
| LV internal dimension in diastole, cm | 88 | 4.3 ± 0.6 | 2.4 ± 1.7 | | 0.2 |
| LV internal dimension in systole, cm | 88 | 2.8 ± 0.4 | 3.8 ± 2.2 | | 0.08 |
| LV ejection fraction, % | 88 | 59.8 ± 3.5 | -0.03 ± 0.3 | | 0.9 |
| LV mass, g | 88 | 113.4 ± 27.8 | 0.02 ± 0.03 | | 0.5 |
| Interventricular septum, cm | 88 | 0.83 ± 0.14 | -10.0 ± 6.9 | | 0.1 |
| LV posterior wall, cm | 88 | 0.84 ± 0.12 | -5.0 ± 7.9 | | 0.5 |

**Table 3B.** Distribution of VTI_QRS-3D_ among healthy males.

| **Variables**<br>**(Healthy males)** | **n** | **VTI<sub>QRS-3D</sub>, μVs**<br>**Mean ± SD** | **IQR** | **Reference ranges**<br>**2.5<sup>th</sup>-97.5<sup>th</sup>**<br>**percentile** | **p-value** |
| --- | --- | --- | --- | --- | --- |
| **Overall** | 169 | 42.3 ± 9.2 | 35.4-49.9 | 25.6-57.2 | - |
| **Age, years** |  |  |  |  | **<0.0001** |
| 18-34 | 58 | 46.5 ± 7.6 | 42.2-51.5 | 29.9-57.2 | |
| 35-54 | 61 | 42.2 ± 8.1 | 35.7-49.6 | 28.2-56.7 | |
| 55-64 | 31 | 37.0 ± 9.4 | 29.2-41.9 | 25.1-56.7 | |
| $\geq 65$ | 19 | 38.4 ± 10.4 | 32.9-45.5 | 18.3-53.9 | |
| **Race** |  |  |  |  | 0.5 |
| White | 111 | 41.8 ± 9.0 | 35.3-49.3 | 26.4-57.3 | |
| Black | 20 | 44.4 ± 9.5 | 39.2-51.2 | 24.0-55.5 | |
| Other/unknown | 38 | 42.8 ± 9.5 | 35.3-50.2 | 25.6-57.0 | |
| **Body Composition** |  | **Mean ± SD** | **β-**<br>**coefficient** |  |  |
| Body surface area, m<sup>2</sup> | 167 | 2.0 ± 0.3 | -4.3 ± 2.4 | | 0.08 |
| Body mass index, kg/m<sup>2</sup> | 167 | 26.5 ± 5.2 | -0.2 ± 0.1 | | 0.08 |
| **Echocardiography** |  | **Mean ± SD** | **β-**<br>**coefficient** |  |  |
| LV internal dimension in diastole, cm | 44 | 4.7 ± 0.6 | 5.0 ± 2.2 | | **0.03** |
| LV internal dimension in systole, cm | 44 | 3.0 ± 0.5 | 4.6 ± 2.8 | | 0.1 |
| LV ejection fraction, % | 44 | 59.1 ± 3.5 | -0.9 ± 0.4 | | **0.01** |
| LV mass, g | 44 | 154.5 ± 42.7 | 0.04 ± 0.03 | | 0.2 |
| Interventricular septum, cm | 44 | 0.96 ± 0.20 | -4.1 ± 6.5 | | 0.5 |
| LV posterior wall, cm | 44 | 0.95 ± 0.16 | 3.6 ± 8.2 | | 0.7 |

Echocardiographic variables were available for 88 (29.4%) females and 44 (26.0%) males. These patients had normal echocardiographic values with small variability. In this relatively homogenous population, the echocardiographic variables exhibited no statistical associations with VTI_QRS-3D_ for females and only had minor statistical significance in males for LVIDd (univariate β =5.0 ± 2.2, p =0.03) and LVEF (univariate β =−0.9 ± 0.4, p =0.01).

As shown in **Table 4**, in the healthy group, the statistically significant multivariate predictors of VTI_QRS-3D_ were age (β =−0.14 ± 0.02, p <0.0001) and female sex (β =−6.41 ± 0.91, p <0.0001).

**Table 4.** Univariate and multivariate predictors of VTI_QRS-3D_ in the overall healthy group.

| Healthy group (n = 468) |  |  |  |  |
| --- | --- | --- | --- | --- |
|  | Univariate |  | Multivariate* |  |
| Variable | β-coefficient | p-value | β-coefficient | p-value |
| Age, years | -0.15 ± 0.02 | **<0.0001** | -0.14 ± 0.02 | **<0.0001** |
| Female | -6.56 ± 0.85 | **<0.0001** | -6.41 ± 0.91 | **<0.0001** |
| Race |  |  |  |  |
| White | Ref | Ref | Ref | Ref |
| Black | 1.79 ± 1.21 | 0.1 | 0.72 ± 1.14 | 0.5 |
| Other | 1.13 ± 1.15 | 0.3 | -0.92 ± 1.09 | 0.4 |
| Body surface area, kg/m<sup>2</sup> | 3.52 ± 1.57 | **0.02** | -0.07 ± 1.61 | 1.0 |

\* Multivariate model with predictors age (linear), female, race and body surface area (linear)

### Cardiomyopathy group

Variables for females and males in the cardiomyopathy group are summarized in **Table 5A and 5B.** In females, VTI_QRS-3D_ was comparable across age groups, with no significant differences noted (p = 0.9). Similarly, values were comparable across racial groups (p = 0.3) and cardiomyopathy types (ischemic vs. non-ischemic, p = 0.3). In males, VTI_QRS-3D_ showed no significant age-related trends (p = 0.2), but there were differences across racial groups (p = 0.007) and between ischemic and non-ischemic cardiomyopathy types (44.3 ± 17.3 µVs vs. 54.4 ± 21.2 µVs, p = 0.0003). Among echocardiographic variables, VTI_QRS-3D_ was positively associated with LV dimensions and calculated LVMi. VTI_QRS-3D_ was negatively associated with LVEF in females (β =−0.7 ± 0.3, p = 0.01) with a similar but weaker trend in males (β =−0.3 ± 0.1, p = 0.08). In the cardiomyopathy group, non-ischemic cardiomyopathy (β =6.74 ± 2.46, p = 0.006) and LVMi (β =0.25 ± 0.03, p <0.0001) were significant multivariate predictors of VTI_QRS-3D_ (**Table 6**).

**Table 5A.**
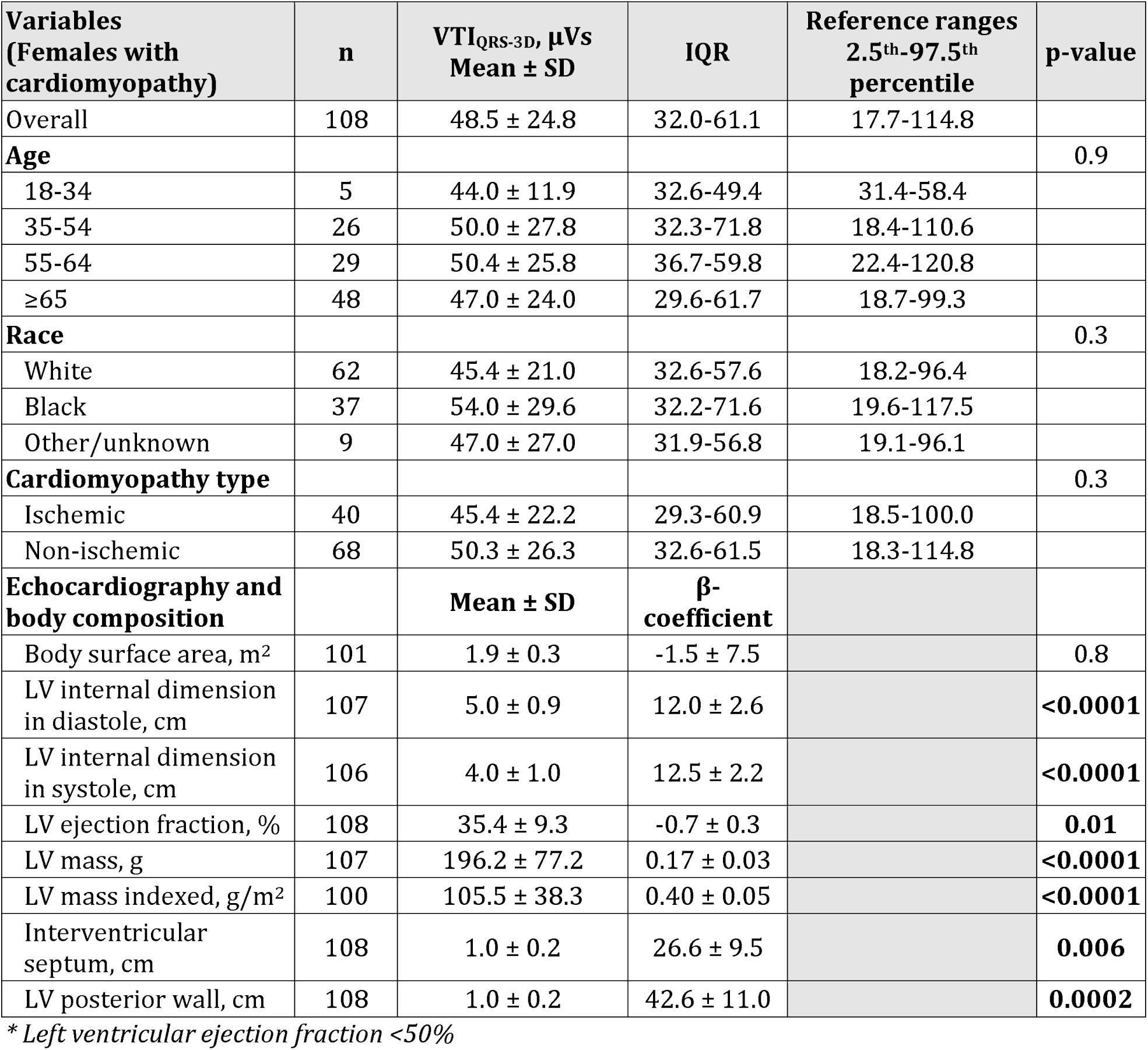
Distribution of VTI_QRS-3D_ among cardiomyopathy with reduced EF* females.

| **Variables**<br>**(Females with**<br>**cardiomyopathy)** | **n** | **VTI<sub>QRS-3D</sub>, μVs**<br>**Mean ± SD** | **IQR** | **Reference ranges**<br>**2.5<sup>th</sup>-97.5<sup>th</sup>**<br>**percentile** | **p-value** |
| --- | --- | --- | --- | --- | --- |
| Overall | 108 | 48.5 ± 24.8 | 32.0-61.1 | 17.7-114.8 | |
| **Age** |  |  |  |  | 0.9 |
| 18-34 | 5 | 44.0 ± 11.9 | 32.6-49.4 | 31.4-58.4 | |
| 35-54 | 26 | 50.0 ± 27.8 | 32.3-71.8 | 18.4-110.6 | |
| 55-64 | 29 | 50.4 ± 25.8 | 36.7-59.8 | 22.4-120.8 | |
| $\geq 65$ | 48 | 47.0 ± 24.0 | 29.6-61.7 | 18.7-99.3 | |
| **Race** |  |  |  |  | 0.3 |
| White | 62 | 45.4 ± 21.0 | 32.6-57.6 | 18.2-96.4 | |
| Black | 37 | 54.0 ± 29.6 | 32.2-71.6 | 19.6-117.5 | |
| Other/unknown | 9 | 47.0 ± 27.0 | 31.9-56.8 | 19.1-96.1 | |
| **Cardiomyopathy type** |  |  |  |  | 0.3 |
| Ischemic | 40 | 45.4 ± 22.2 | 29.3-60.9 | 18.5-100.0 | |
| Non-ischemic | 68 | 50.3 ± 26.3 | 32.6-61.5 | 18.3-114.8 | |
| **Echocardiography and**<br>**body composition** |  | **Mean ± SD** | **β-**<br>**coefficient** |  |  |
| Body surface area, m<sup>2</sup> | 101 | 1.9 ± 0.3 | -1.5 ± 7.5 | | 0.8 |
| LV internal dimension<br>in diastole, cm | 107 | 5.0 ± 0.9 | 12.0 ± 2.6 | | <0.0001 |
| LV internal dimension<br>in systole, cm | 106 | 4.0 ± 1.0 | 12.5 ± 2.2 | | <0.0001 |
| LV ejection fraction, % | 108 | 35.4 ± 9.3 | -0.7 ± 0.3 | | 0.01 |
| LV mass, g | 107 | 196.2 ± 77.2 | 0.17 ± 0.03 | | <0.0001 |
| LV mass indexed, g/m<sup>2</sup> | 100 | 105.5 ± 38.3 | 0.40 ± 0.05 | | <0.0001 |
| Interventricular<br>septum, cm | 108 | 1.0 ± 0.2 | 26.6 ± 9.5 | | 0.006 |
| LV posterior wall, cm | 108 | 1.0 ± 0.2 | 42.6 ± 11.0 | | 0.0002 |

\* Left ventricular ejection fraction <50%

**Table 5B.** Distribution of VTI_QRS-3D_ among cardiomyopathy with reduced EF* males.

| **Variables**<br>**(Males with**<br>**cardiomyopathy)** | **n** | **VTI<sub>QRS-3D</sub>, μVs**<br>**Mean ± SD** | **IQR** | **Reference ranges**<br>**2.5<sup>th</sup>-97.5<sup>th</sup>**<br>**percentile** | **p-value** |
| --- | --- | --- | --- | --- | --- |
| **Overall** | 206 | 48.1 ± 19.4 | 33.1-58.8 | 21.7-98.3 | |
| **Age, years** |  |  |  |  | 0.2 |
| 18-34 | 9 | 61.9 ± 35.5 | 40.1-77.3 | 25.7-122.6 | |
| 35-54 | 54 | 49.1 ± 21.5 | 33.3-61.4 | 21.2-86.8 | |
| 55-64 | 54 | 46.9 ± 16.7 | 33.0-56.6 | 23.1-84.7 | |
| $\geq 65$ | 89 | 46.9 ± 17.2 | 33.7-55.5 | 22.7-93.4 | |
| **Race** |  |  |  |  | **0.007** |
| White | 135 | 45.0 ± 16.5 | 32.7-54.4 | 21.8-86.7 | |
| Black | 42 | 53.7 ± 18.4 | 41.6-62.1 | 24.7-108.7 | |
| Other/unknown | 29 | 54.3 ± 28.7 | 33.2-63.9 | 22.3-129.7 | |
| **Cardiomyopathy type** |  |  |  |  | **0.0003** |
| Ischemic | 128 | 44.3 ± 17.3 | 32.4-50.5 | 21.7-86.9 | |
| Non-ischemic | 78 | 54.4 ± 21.2 | 40.1-63.4 | 25.4-110.6 | |
| **Echocardiography and**<br>**body composition** |  | **Mean ± SD** | **β-**<br>**coefficient** |  |  |
| Body surface area, m<sup>2</sup> | 190 | 2.0 ± 0.3 | -4.2 ± 5.1 | | 0.4 |
| LV internal dimension in<br>diastole, cm | 204 | 5.4 ± 0.8 | 4.5 ± 1.8 | | **0.01** |
| LV internal dimension in<br>systole, cm | 203 | 4.2 ± 0.9 | 6.3 ± 1.4 | | **<0.0001** |
| LV ejection fraction, % | 206 | 36.2 ± 9.5 | -0.3 ± 0.1 | | 0.08 |
| LV mass, g | 204 | 231.9 ± 78.7 | 0.08 ± 0.02 | | **<0.0001** |
| LV mass indexed, g/m<sup>2</sup> | 189 | 113.3 ± 34.7 | 0.20 ± 0.04 | | **<0.0001** |
| Interventricular septum,<br>cm | 204 | 1.1 ± 0.2 | 12.5 ± 5.7 | | **0.03** |
| LV posterior wall, cm | 204 | 1.1 ± 0.2 | 20.6 ± 5.8 | | **0.0005** |

\* Left ventricular ejection fraction <50%

**Table 6.** Univariate and multivariate predictors of VTI_QRS-3D_ in the cardiomyopathy* group.

| **Cardiomyopathy with reduced EF group (n = 314)** |  |  |  |  |
| --- | --- | --- | --- | --- |
| **Variable** | **Univariate** |  | **Multivariate** |  |
| | β-coefficient | p-value | β-coefficient | p-value |
| Age | $-0.02 \pm 0.09$ | 0.8 | $-0.03 \pm 0.09$ | 0.8 |
| Female | $0.40 \pm 2.55$ | 0.9 | $0.23 \pm 2.58$ | 0.9 |
| Race |  |  |  |  |
| White | Ref | Ref | Ref | Ref |
| Black | $8.69 \pm 2.81$ | **0.002** | $5.01 \pm 2.73$ | 0.07 |
| Other | $7.42 \pm 3.73$ | **0.048** | $5.68 \pm 3.58$ | 0.1 |
| Body surface area | $-2.98 \pm 4.06$ | 0.5 | $-2.47 \pm 3.95$ | 0.5 |
| Type of cardiomyopathy |  |  |  |  |
| Ischemic | Ref | Ref | Ref | Ref |
| Non-ischemic | $7.92 \pm 2.38$ | **0.001** | $6.74 \pm 2.46$ | **0.006** |
| LV ejection fraction (%) | $-0.39 \pm 0.13$ | **0.003** | $-0.08 \pm 0.13$ | 0.5 |
| LV mass indexed | $0.27 \pm 0.03$ | **<0.0001** | $0.25 \pm 0.03$ | **<0.0001** |

\* Left ventricular ejection fraction <50%

### Reference ranges

The overall reference range (2.5th–97.5th percentiles) for the entire healthy population is 20.9–56.4 µVs. The reference range of VTI_QRS-3D_ specifically for females is 20.2–55.7 µVs and for males is 25.6–57.2 µVs. The percentile values of VTI_QRS-3D_ among healthy group for various age groups and sex are shown in **Table 7**. The reference ranges by age groups for females were 23.2–55.0 µVs (18–34 years), 23.9–56.4 µVs (35–54 years), and 19.6–50.9 µVs (≥55 years). For males, the ranges were 29.9–57.2 µVs, 28.2–56.7 µVs, and 21.4–55.9 µVs, respectively.

**Table 7.**
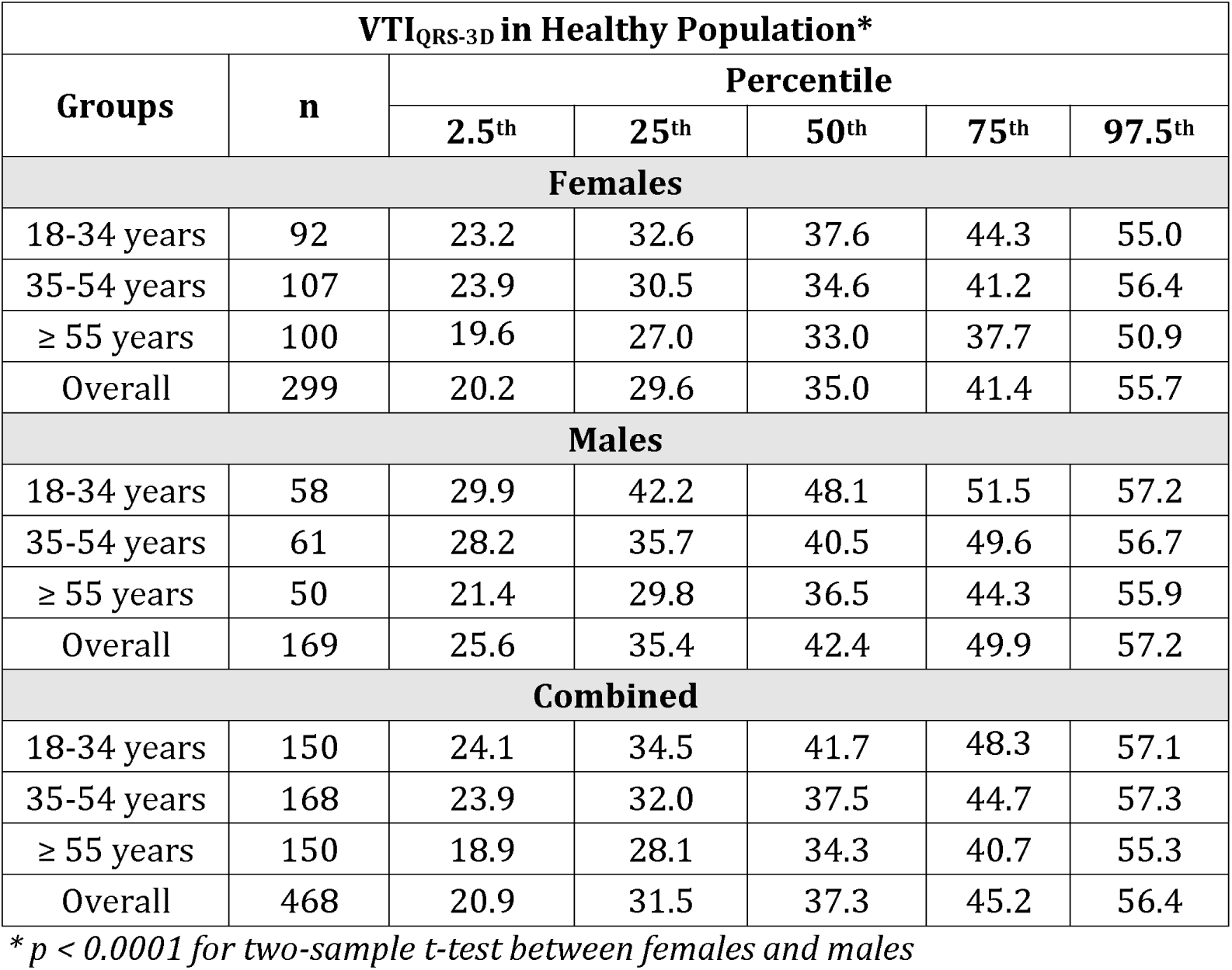
Reference ranges of VTI_QRS-3D_ in healthy group (N=468)

## DISCUSSION

In this study, we computed the reference values for VTI_QRS-3D_, an automatically calculable measurement with potential for integration into automated ECG analysis. Several features of VTI_QRS-3D_, including robust automated calculation and efficient summarization of myocardial depolarization in one numerical value, make it a suitable metric for ECG-based research and broader assessment of clinical applications. As opposed to the QRS duration, which has high interobserver and interobserver variability in measurement, the VTI_QRS-3D_ is very reproducible as it weights the QRS duration to the 3D/RMS voltage at any instant during the QRS, thereby assigning very small weights to the beginning and end of QRS. On the other hand, as opposed to QRS voltage alone, VTI does incorporate the QRS duration and is therefore a more accurate representation of the summed force of the ventricular activation ECG potential. The average VTI_QRS-3D_ in our healthy group was 38.1 ± 9.3 µVs and among cardiomyopathy with reduced EF patients was 48.2 ± 21.4 µVs.

### Terminology disambiguation

The literature contains various terms to describe VTI_QRS-3D_ and related measurements. One commonly used metric is 3D QRS area (or QRS_AREA_), which is derived by calculating the root-mean-square of the individual voltage-time integrals of QRS projections along the X, Y and Z axes, and has been used in studies evaluating CRT response.^3,13^ 3D QRS area differs from VTI_QRS-3D_, where voltage-time integral is calculated from the scalar 3D lead obtained by plotting root-mean-square of the X, Y and Z leads. Further, 3D QRS area can be calculated using two methods: the summation method and the difference method. In the summation method, areas under the positive and negative deflections along each lead are added, while in the difference method, they are subtracted. We have shown previously that the values of 3D QRS area obtained through the summation method are close to the VTI_QRS-3D_ values in normal ECGs (linear regression, β 1.07, R^2^ 0.99), while those obtained using the difference method can diverge significantly (β 1.42, R^2^ 0.65).^14^ In older studies, spatial vector of QRS (SÂ QRS) has been used by Pipberger et al. to describe 3D QRS area obtained via the difference method.^15,16^ Of note, they also included P-wave integrals in this metric, assuming the P-wave contribution to be negligible. Tereshchenko et al. introduced the sum absolute QRST integral (SAI QRST), which is calculated using the arithmetic sum of orthogonal lead areas (derived via the summation method) instead of the root mean square used for the 3D QRS area and includes the T-wave in its computation.^17^ Later, the term SAI QRS has been used as well, which does not incorporate the T-wave.^18^ Although these interrelated metrics differ in their calculation, their general associations with clinical covariates are expected to remain similar.

### Trends in VTI _QRS-3D_

We noted several trends of VTI_QRS-3D_ with covariates. Foremost, in the healthy group, older age and female sex were associated with smaller values of VTI_QRS-3D_. Notably, age stratification revealed that the negative correlation between age and VTI_QRS-3D_ persists up to approximately 65 years of age, beyond which it stabilizes or may even show a slight increase. A decrease in SAI QRS and SÂ QRS with age in healthy subjects has been observed in previous studies as well.^15,18^ The mechanism for this remains unclear although this may be attributable to cardiac atrophy that occurs with age.^19^ Beyond 65 years, increased incidence of asymptomatic structural heart disease and conduction abnormalities may explain the stabilization or increase noted in VTI_QRS-3D_ values.. Further, it is widely known that the cardiac dimensions and measured ECG voltages are smaller in females as compared to males, and this is reflected in the values of VTI_QRS-3D_.^7^

Second, patients having cardiomyopathy with reduced EF had significantly larger VTI_QRS-3D_ as compared to healthy patients. This is an expected finding, since cardiomyopathy with reduced EF is associated with ventricular activation delay and increased LV mass/volume, which may lead to prolonged QRS duration and increased QRS voltage.^20–22^ Previous studies show that cardiac resynchronization therapy (CRT) response is better in patients with larger 3D QRS area, since it reflects delayed LV activation which can be mitigated by CRT.^3^ Furthermore, VTI_QRS-3D_ increases with LV mass and serves as a superior ECG predictor of left ventricular hypertrophy, when compared to previously published voltage-based criteria.^7,23^

Third, in the cardiomyopathy with reduced EF group, patients with non-ischemic as opposed to ischemic cardiomyopathy and higher left ventricular mass indexed (LVMi) had larger VTI_QRS-3D_. The association between nonischemic cardiomyopathy and increased 3D QRS area has also been observed in multiple previous studies.^13,24,25^ Interestingly, unlike in the healthy group, age and sex did not show statistically significant associations with VTI_QRS-3D_ in patients having cardiomyopathy with reduced EF. This suggests that specific characteristics of cardiomyopathy, such as underlying mechanisms and increased LV mass, may serve as dominant drivers or effect modifiers, obfuscating the effect of age and sex on VTI_QRS-3D_ in this population. However, given that the cardiomyopathy group in our dataset was predominantly composed of older patients (70% aged ≥55 years), the imbalance in age distribution may have limited the ability to detect a statistically significant association between age and VTI_QRS-3D_.

### Previously published reference ranges

Pipberger et al. published age-based reference ranges of SÂ QRS for 518 normal men in 1967.^15^ Similar to our results, they observed a decline in SÂ QRS with age, with mean values ranging from 42.0 ± 13.0 µVs in 20-29 years group to 32.4 ± 13.4 µVs in the 60-78 years group. In our sample, the mean VTI_QRS-3D_ values in healthy men were shifted higher, from 46.5 ± 7.6 µVs in 18-34 years group to 38.4 ± 10.4 µVs in ≥65 years group. The differences in Pipberger et al. and our magnitudes may be accounted by the differences in the calculation of SÂ QRS versus VTI_QRS-3D_. SÂ QRS of Pipberger et al. is equivalent to 3D QRS area calculated using the difference method, which is a systematic underestimate of VTI_QRS-3D_.^14^ Further, Pipberger et al. used the original Frank orthogonal lead system to record vectorcardiograms, whereas we derived orthogonal leads from standard 12-lead ECGs using the Kors matrix.^26^

More recently, in a comprehensive analysis of various vectorcardiography parameters, De la Garza Salazar and Egenriether studied mean values of SAI QRS for different categories of age, sex, BMI, hypertension, ischemic heart disease, and left ventricular hypertrophy.^18^ Owing to differences in calculation of these metrics (we took the time integral of instantaneous RMS voltage while they took the arithmetic sum of the integrals in X, Y and Z leads), we observed overall mean VTI_QRS-3D_ value of approximately 40 µVs, whereas they observed mean SAI QRS values closer to 60 µVs across groups. Similar to our results, they observed a decrease in SAI QRS values with increased age and female sex.

### Strengths and limitations

The main strength of our analysis is the delineation of a healthy population verified through a manual chart review, ensuring that ECGs in the healthy group belong to patients without pre-existing cardiovascular disease. Our analysis encompassed a diverse population including various ages, sexes and races. However, there are several important limitations to our analysis as well. These include a limited sample size, limited racial and ethnic diversity, and the use of clinical ECGs rather than seeking ECGs from healthy volunteers in the community.

## CONCLUSIONS

Values of VTI_QRS-3D_ are higher at younger age in healthy population, male sex and in patients having cardiomyopathy with reduced EF. If adopted for future clinical reporting, VTI_QRS-3D_ reference ranges should be interpreted in the context of these predictors. Further, there is a need to standardize terminology and computation algorithms for VTI_QRS-3D_ and related QRS area metrics.

## Data Availability

The data supporting the findings of this study were obtained from an institutional database that includes identifiable patient information. Due to privacy and ethical considerations, these data cannot be shared publicly. Access to the data is restricted and subject to approval by the institutional review board (IRB). Researchers interested in accessing the data may contact the corresponding author or the institution's data governance committee for information about the necessary procedures and approvals required.

## Project support

This project was supported by the NIH CTSA award # UL1TR002366

## REFERENCES

1. Frank E. An Accurate, Clinically Practical System For Spatial Vectorcardiography. Circulation. 1956;13:737–749. doi: 10.1161/01.CIR.13.5.737

2. Jaros R, Martinek R, Danys L. Comparison of Different Electrocardiography with Vectorcardiography Transformations. Sensors (Basel). 2019;19. doi: 10.3390/s19143072

3. van Stipdonk AMW, Ter Horst I, Kloosterman M, Engels EB, Rienstra M, Crijns H, Vos MA, van Gelder IC, Prinzen FW, Meine M, et al. QRS Area Is a Strong Determinant of Outcome in Cardiac Resynchronization Therapy. Circ Arrhythm Electrophysiol. 2018;11:e006497. doi: 10.1161/CIRCEP.118.006497

4. Morey T, Harvey Christopher J, Mahmood U, Parimi N, Lacy S, DeBauge A, Sheldon S, Reddy M, Noheria A. CHANGE IN QRS 3D VOLTAGE TIME INTEGRAL (3D QRS AREA) WITH CARDIAC RESYNCHRONIZATION THERAPY PREDICTS SUBSEQUENT LEFT VENTRICULAR REVERSE REMODELING AND HEART FAILURE HOSPITALIZATIONS. Journal of the American College of Cardiology. 2021;77:359–359. doi: 10.1016/S0735-1097(21)01718-6

5. Noheria A, Sodhi S, Orme GJ. The Evolving Role of Electrocardiography in Cardiac Resynchronization Therapy. Curr Treat Options Cardiovasc Med. 2019;21:91. doi: 10.1007/s11936-019-0784-6

6. Bank AJ, Brown CD, Burns KV, Espinosa EA, Harbin MM. Electrical dyssynchrony mapping and cardiac resynchronization therapy. J Electrocardiol. 2022;74:73–81. doi: 10.1016/j.jelectrocard.2022.08.006

7. DeBauge A, Harvey CJ, Gupta A, Fairbank T, Ranka S, Jiwani S, Reddy M, Sheldon SH, Noheria A. Evaluation of electrocardiographic criteria for predicting left ventricular hypertrophy and dilation in presence of left bundle branch block. J Electrocardiol. 2024;87:153787. doi: 10.1016/j.jelectrocard.2024.153787

8. DeBauge A, Fairbank T, Harvey CJ, Ranka S, Jiwani S, Sheldon SH, Reddy M, Beaver TA, Noheria A. Electrocardiographic prediction of left ventricular hypertrophy in women and men with left bundle branch block - Comparison of QRS duration, amplitude and voltage-time-integral. J Electrocardiol. 2023;80:34–39. doi: 10.1016/j.jelectrocard.2023.03.004

9. Waitman LR, Warren JJ, Manos EL, Connolly DW. Expressing observations from electronic medical record flowsheets in an i2b2 based clinical data repository to support research and quality improvement. AMIA Annu Symp Proc. 2011;2011:1454–1463.

10. Murphy SN, Weber G, Mendis M, Gainer V, Chueh HC, Churchill S, Kohane I. Serving the enterprise and beyond with informatics for integrating biology and the bedside (i2b2). J Am Med Inform Assoc. 2010;17:124–130. doi: 10.1136/jamia.2009.000893

11. Lang RM, Bierig M, Devereux RB, Flachskampf FA, Foster E, Pellikka PA, Picard MH, Roman MJ, Seward J, Shanewise J, et al. Recommendations for chamber quantification. Eur J Echocardiogr. 2006;7:79–108. doi: 10.1016/j.euje.2005.12.014

12. Kors JA, van Herpen G, Sittig AC, van Bemmel JH. Reconstruction of the Frank vectorcardiogram from standard electrocardiographic leads: diagnostic comparison of different methods. Eur Heart J. 1990;11:1083–1092. doi: 10.1093/oxfordjournals.eurheartj.a059647

13. van Deursen CJ, Vernooy K, Dudink E, Bergfeldt L, Crijns HJ, Prinzen FW, Wecke L. Vectorcardiographic QRS area as a novel predictor of response to cardiac resynchronization therapy. J Electrocardiol. 2015;48:45–52. doi: 10.1016/j.jelectrocard.2014.10.003

14. Noheria A, Toquica C, Mahmood UA, DeBauge A, Morey T, Harvey CJ. Different methods of 3D QRS area calculation from vectorcardiographic X, Y, and Z Leads. Pacing Clin Electrophysiol. 2024;47:974–976. doi: 10.1111/pace.14968

15. Pipberger HV, Goldman MJ, Littmann D, Murphy GP, Cosma J, Snyder JR. Correlations of the orthogonal electrocardiogram and vectorcardiogram with consitutional variables in 518 normal men. Circulation. 1967;35:536–551. doi: 10.1161/01.cir.35.3.536

16. Draper HW, Peffer CJ, Stallmann FW, Littmann D, Pipberger HV. The Corrected Orthogonal Electrocardiogram and Vectorcardiogram in 510 Normal Men (Frank Lead System). Circulation. 1964;30:853–864. doi: 10.1161/01.cir.30.6.853

17. Tereshchenko LG, Cheng A, Fetics BJ, Marine JE, Spragg DD, Sinha S, Calkins H, Tomaselli GF, Berger RD. Ventricular arrhythmia is predicted by sum absolute QRST integralbut not by QRS width. J Electrocardiol. 2010;43:548–552. doi: 10.1016/j.jelectrocard.2010.07.013

18. De la Garza Salazar F, Egenriether B. Exploring vectorcardiography: An extensive vectocardiogram analysis across age, sex, BMI, and cardiac conditions. J Electrocardiol. 2024;82:100–112. doi: 10.1016/j.jelectrocard.2023.12.004

19. Sur S, Han L, Tereshchenko LG. Comparison of sum absolute QRST integral, and temporal variability in depolarization and repolarization, measured by dynamic vectorcardiography approach, in healthy men and women. PLoS One. 2013;8:e57175. doi: 10.1371/journal.pone.0057175

20. Andersen DC, Kragholm K, Petersen LT, Graff C, Sorensen PL, Nielsen JB, Pietersen A, Sogaard P, Atwater BD, Friedman DJ, et al. Association between vectorcardiographic QRS area and incident heart failure diagnosis and mortality among patients with left bundle branch block: A register-based cohort study. J Electrocardiol. 2021;69:30–35. doi: 10.1016/j.jelectrocard.2021.09.002

21. Aimo A, Milandri A, Barison A, Pezzato A, Morfino P, Vergaro G, Merlo M, Argiro A, Olivotto I, Emdin M, et al. Electrocardiographic abnormalities in patients with cardiomyopathies. Heart Fail Rev. 2024;29:151–164. doi: 10.1007/s10741-023-10358-7

22. Mafi Rad M, Wijntjens GW, Engels EB, Blaauw Y, Luermans JG, Pison L, Crijns HJ, Prinzen FW, Vernooy K. Vectorcardiographic QRS area identifies delayed left ventricular lateral wall activation determined by electroanatomic mapping in candidates for cardiac resynchronization therapy. Heart Rhythm. 2016;13:217–225. doi: 10.1016/j.hrthm.2015.07.033

23. DeBauge A, Harvey C, Gupta A, Noheria A. Abstract 4137204: Classifying Left Ventricular Hypertrophy from ECG in Overall Population and Bundle Branch Blocks: Machine Learning Models are Superior to Published ECG Criteria. Circulation. 2024;150:A4137204–A4137204. doi: 10.1161/circ.150.suppl_1.4137204

24. Okafor O, Zegard A, van Dam P, Stegemann B, Qiu T, Marshall H, Leyva F. Changes in QRS Area and QRS Duration After Cardiac Resynchronization Therapy Predict Cardiac Mortality, Heart Failure Hospitalizations, and Ventricular Arrhythmias. J Am Heart Assoc. 2019;8:e013539. doi: 10.1161/JAHA.119.013539

25. Nguyen UC, Claridge S, Vernooy K, Engels EB, Razavi R, Rinaldi CA, Chen Z, Prinzen FW. Relationship between vectorcardiographic QRS(area), myocardial scar quantification, and response to cardiac resynchronization therapy. J Electrocardiol. 2018;51:457–463. doi: 10.1016/j.jelectrocard.2018.01.009

26. Frank E. An accurate, clinically practical system for spatial vectorcardiography. Circulation. 1956;13:737–749. doi: 10.1161/01.cir.13.5.737

